# Prevalence of skin disorders and associated factors among patients with celiac disease: a cross-sectional study

**DOI:** 10.1101/2025.01.12.25320431

**Authors:** Zohreh Tajabadi, Golnaz Ekhlasi, Maryam Daneshpazhooh, Hafez Fakheri, Tahmine Tavakoli, Bizhan Ahmadi, Azita Ganji, Ramin Niknam, Mohammad Reza Pashaei, Mohammad Reza Ghadir, Shaahin Shahbazi, Fariborz Mansour Ghanaei, Pezhman Alavinejad, Masoud Shirmohamadi, Majid Aflatoonian, Alireza Bakhshipour, Sahar Masoudi, Bijan Shahbazkhani

## Abstract

**Background:** While celiac disease (CD) ordinarily presents with gastrointestinal manifestations, extraintestinal appearances may also happen. Although cutaneous manifestations are one of the most common extraintestinal manifestations of CD, little is known about their prevalence and associated factors. This study aims to determine the prevalence of cutaneous manifestations and related factors in CD patients.

**Methods:** This cross-sectional study enrolled CD patients referred to the National Celiac Registry, for whom a diagnosis of CD was confirmed through clinical examination, serological testing, and intestinal biopsy. Data on demographic characteristics, CD staging, clinical manifestations, underlying comorbidities, and family history (FH) of diseases were collected. Data were analyzed using SPSS version 25.0. A *p*-value less than 0.05 was considered significant.

**Results:** Of the 4357 enrolled CD patients (male 65.7%), 18.20% had cutaneous manifestations, with eczema (3.73%), dermatitis herpetiformis (3.46%), and psoriasis (0.71%) being the most common disorders. The FH of skin disorders (Odds ratio (OR)=7.99, 95% confidence interval (CI) 2.44-26.13, *p*=0.00), osteoarthritis (OR=6.15, 95% CI 1.15-32.78, *p*=0.03), Sjögren syndrome (OR=4.82, 95% CI 1.98-11.73, *p*=0.00), mouth aphthae (OR=3.10, 95% CI 1.80-5.32, *p*=0.00), thalassemia (OR=2.78, 95% CI 1.19-6.50, *p*=0.01), constipation (OR =1.62, 95% CI 1.10-2.38, *p*=0.01), and iron deficiency anemia (IDA) (OR=1.54, 95% CI 1.05-2.24, *p*=0.02) were independent predictors, and type I diabetes mellitus (OR=0.20, 95% CI 0.05-0.81, *p*=0.02) was a protective factor for skin diseases.

**Conclusion:** Considering the higher probability of cutaneous manifestations in CD patients, regular clinical evaluation of these patients is recommended for timely diagnosis and management of underlying skin diseases.

## Introduction

Celiac disease (CD) is a chronic, immune-mediated disorder triggered by exposure to dietary gluten in genetically susceptible individuals (1). The prevalence of the disease is up to 1% worldwide (2) and varies among different geographical regions, ranging from 0.2% in Germany to 5.6% in Africa. In recent years, the incidence and prevalence of CD have been on the rise (3, 4), partly due to increased consumption of gluten-containing products and improved accessibility to serological tests with high specificity and sensitivity (5).

CD primarily affects the small intestine and may manifest as an asymptomatic disease or present with typical gastrointestinal manifestations (6). Additionally, atypical extra-intestinal manifestations may also occur (7), and in some cases, such manifestations could be the sole presentation of CD. Certain individuals with CD may exhibit various cutaneous manifestations. A large study carried out by Lebwohl et al. indicated that celiac patients had a significantly higher risk of developing skin manifestations compared to healthy individuals (8). Besides dermatitis herpetiformis (DH), which is considered as the most common cutaneous manifestation of CD, the prevalence of several other skin disorders including psoriasis, eczema, and urticaria, has been found to be higher among these patients compared to healthy individuals (8, 9). However, the exact association between CD and skin disorders still remains inconclusive.

Celiac autoantibodies have been detected in patients with skin disorders like DH (10). Furthermore, improvement in clinical manifestations has been observed in patients with skin disorders after receiving a gluten-free diet (GFD) (11). Studies also showed that skin disorders might be associated with various autoimmune (AI) diseases such as type I diabetes mellitus (DM) and thyroiditis, which are also common among celiac patients (12, 13). In addition, several shared genetic loci have been found between CD and skin disorders such as DH (14) and psoriasis (15). Furthermore, the immune system may also play an important role in the pathogenesis of both CD and skin disorders. For instance, an increased number of T cells and T cell-mediated inflammation were found among patients with psoriasis and CD (16, 17). Altogether, these findings may suggest a possible linkage between CD and skin disorders. However, the exact underlying mechanisms are yet to be understood (18).

Despite the high prevalence of CD and associated skin disorders in Middle Eastern countries such as Iran (19), previous studies in this region have not comprehensively evaluated CD patients with cutaneous disorders. Understanding the interplay between CD and skin disorders, along with associated factors, may lead to early diagnosis and appropriate treatment for affected patients. Therefore, the present study aims to evaluate the prevalence of cutaneous disorders in CD patients and assess related demographic, clinical, and histopathological factors which may affect the probability of developing cutaneous manifestations in affected patients.

## Methods

### Study setting and population

This cross-sectional study aims to evaluate the prevalence of skin disorders among CD patients and compare the demographic, clinical, and histopathological characteristics of affected patients with and without cutaneous manifestations. The study recruited patients referred to the Celiac Registry Center in Shariati Hospital, Tehran University of Medical Sciences, Tehran, Iran, between 2019 and 2022. The diagnosis of CD was confirmed by an expert gastroenterologist through clinical examination, intestinal histopathological study, and serological evaluation. Patients meeting the inclusion criteria included those with a confirmed diagnosis of CD based on intestinal biopsy and serological examination, and who had willingness to participate in the study. Exclusion criteria comprised individuals with Marsh<2, negative anti-tissue transglutaminase (tTG) test result, inflammatory bowel disease, lack of willingness to participate, and incomplete information.

### CD diagnosis protocol

The diagnosis of CD involved clinical examination, serological evaluation, and intestinal histopathological study. Serum levels of anti-tTG IgA and IgG antibodies were evaluated using the ELISA method in patients displaying suspicious clinical manifestations. Those with positive anti-tTG test results underwent gastrointestinal endoscopy and biopsy by an expert gastroenterologist. Four to six samples were taken from the small intestine and evaluated by pathologists. The severity of intestinal villous atrophy was graded based on the Marsh staging system, in which Marsh 0 means normal villous and Marsh IIIc is considered as total villous atrophy. Subjects with positive anti-tTG test results and Marsh II or higher were diagnosed with CD.

### Data collection

Data on the patient’s age, gender, gastrointestinal, cutaneous, oro-dental, hematological, and neurological manifestations, underlying medical conditions, and family history (FH) of associated disorders were collected through interviews conducted by a trained interviewer. The diagnosis of skin disorders was made by a dermatologist using clinical manifestations and histopathological examination if needed. Information on Marsh staging was obtained from pathological reports, while data on underlying disorders was extracted from patient medical records. The collected data were entered into an electronic database and retrospectively analyzed (access date 10^th^ December 2023). The researchers were blind to participant identity. The primary objective of the study was to determine the prevalence of cutaneous disorders and compare the demographic, clinical, and histopathological characteristics between CD patients with and without skin disorders. The secondary objective involved examining the association between the aforementioned variables and risk of developing skin disorders.

### Ethical issue

The present study was conducted in accordance with the declaration of Helsinki Principles and approved by the Local Ethics Committee of Tehran University of Medical Sciences (IR.TUMS.DDRI.REC.1403.038). Written informed consent was signed by all patients.

### Statistical analysis

Data were analyzed using IBM SPSS Statistics for Windows, version 25.0 (IBM Corp., Armonk, N.Y., USA). Proportions were compared using the Chi-square test and Fisher’s exact test as appropriate. Descriptive statistics were presented as frequencies, percentages, means, and standard deviations (SD). Multivariate logistic regression was performed to evaluate the association between patient age, gender, symptoms, underlying comorbidities, and FH of associated disorders and skin disorders. A *p*-value of less than 0.05 was considered statistically significant.

## Results

### Demographic and baseline characteristics

A total of 4,357 CD patients met the inclusion criteria and were enrolled, 65.7% were male, 81.8% revealed no skin manifestations, and 18.2% reported cutaneous presentations. The proportions of male participants were 67.8% and 65.9% for those without and with cutaneous manifestations, respectively. No statistically significant difference was found in terms of gender between the two study groups (*p*=0.29). The mean age of patients with and without skin manifestations was 29±17 and 27±17 years, respectively, showing a statistically significant difference between the two groups (*p*=0.00). Regarding CD severity, intestinal biopsy revealed that 36.5%, 34.2%, and 29.2% of participants had Marsh IIIa, Marsh IIIb, and Marsh IIIc classifications, respectively. No statistically significant difference was found in Marsh staging between the two groups. Detailed participant characteristics are presented in **Table 1**.

**Table 1.** Demographic and histopathological characteristics of patients with and without cutaneous manifestations.

| Variables | Patient without cutaneous manifestations (n= 3564) | Patients with cutaneous manifestations (n=793) |  |  |  |  | P-value |
| --- | --- | --- | --- | --- | --- | --- | --- |
|  |  | Total (n=793) | Eczema (n=251) | DH (n=179) | Psoriasis (n=47) | Others (n=423) |  |
| Gender, n (%) |  |  |  |  |  |  |  |
| Male | 2347 (67.89) | 515 (65.94) | 160 (64.78) | 112 (64.37) | 30 (63.83) | 286 (68.26) | 0.293 |
| Age at diagnosis, year (mean ± SD) | 27±17 | 29±17 | 27±18 | 31±17 | 33±14 | 29±17 | 0.001 |
| Marsh classification, n (%) |  |  |  |  |  |  |  |
| Marsh IIIa | 1306 (36.64) | 286 (36.07) | 98 (39.04) | 72 (40.22) | 17 (36.17) | 138 (32.62) | 0.819 |
| Marsh IIIb | 1212 (34.01) | 279 (35.18) | 79 (31.47) | 58 (32.40) | 12 (25.53) | 162 (38.30) |  |
| Marsh IIIc | 1046 (29.35) | 228 (28.75) | 74 (29.48) | 49 (27.37) | 18 (38.30) | 123<br>(29.08) |  |

**Abbreviations:** DH, dermatitis herpetiformis; N, number; SD, standard deviation.

### Prevalence of skin disorders

Eczema (3.7%) was the most common cutaneous disease, followed by DH (3.4%) and psoriasis (0.7%). Among the patients, 2.9% had multiple cutaneous manifestations and 7.8% had other types of skin disorders. Among CD patients with cutaneous diseases, 31.6%, 22.5%, 5.9%, and 53.3% were noted to have eczema, DH, psoriasis, and other types of skin disorders, respectively. Details of demographic characteristics of CD patients with each type of skin disorder are shown in **Table 1**.

### Gastrointestinal manifestations

The most common gastrointestinal manifestation was abdominal pain (60.6%), followed by flatulence (50.9%), anorexia (38.7%), diarrhea (33.7%), reflux (31.4%), constipation (30.7%), nausea (26.6%), and vomiting (14.3%). The prevalence of gastrointestinal manifestations was significantly higher among celiac patients with cutaneous disorders compared with those without skin manifestations (*p*<0.05) (**Table 2**).

**Table 2.** Gastrointestinal and extra-intestinal manifestations of patients with and without cutaneous manifestations.

| Variables | Patients<br>without<br>cutaneous<br>manifestations<br>(n= 3564) | Patients with cutaneous manifestations (n=793) |  |  |  |  | P-value |
| --- | --- | --- | --- | --- | --- | --- | --- |
|  |  | Total | Eczema | DH | Psoriasis | Others |  |
|  |  | (n=793) | (n=251) | (n=179) | (n=47) | (n=423) |  |
| Gastrointestinal manifestations, n (%) |  |  |  |  |  |  |  |
| Abdominal pain | 2087 (66.34) | 555 | 190 | 129 | 38 | 292 | **0.012** |
|  |  | (71.06) | (76.61) | (74.14) | (88.37) | (69.19) |  |
| Flatulence | 1703 (55.96) | 518 | 163 | 105 | 35 | 286 | **<0.001** |
|  |  | (67.36) | (67.08) | (62.13) | (81.40) | (68.59) |  |
| Anorexia | 1306 (43.52) | 382 | 133 | 84 | 21 | 199 | **0.001** |
|  |  | (50.33) | (55.65) | (51.22) | (53.85) | (47.72) |  |
| Diarrhea | 1118 (36.97) | 352 | 112 | 74 | 23 | 188 | **<0.001** |
|  |  | (46.19) | (46.47) | (45.12) | (56.10) | (45.08) |  |
| Reflux | 1048 (35.45) | 322 | 90 | 63 | 24 | 191 | **<0.001** |
|  |  | (43.22) | (38.30) | (40.38) | (60.00) | (46.14) |  |
| Constipation | 1010 (33.61) | 329 | 111 | 78 | 22 | 168 | **<0.001** |
|  |  | (43.23) | (45.68) | (47.85) | (52.38) | (40.29) |  |
| Nausea | 884 (29.42) | 274<br>(36.01) | 81<br>(33.47) | 55<br>(33.33) | 15<br>(37.50) | 163<br>(39.18) | **<0.001** |
| Vomiting | 477 (16.17) | 148<br>(19.58) | 51<br>(21.25) | 34<br>(20.86) | 9 (22.50) | 78<br>(18.84) | **0.026** |
| **Oro-dental manifestations, n (%)** |  |  |  |  |  |  |  |
| Mouth aphtae | 377 (21.44) | 163<br>(30.99) | 60<br>(31.91) | 33<br>(25.58) | 12<br>(37.50) | 84<br>(30.77) | **<0.001** |
| Colorless teeth | 160 (9.69) | 35 (7.48) | 21<br>(12.21) | 13<br>(11.71) | 4 (12.50) | 13 (5.42) | 0.144 |
| Mouth ulcer | 103 (6.35) | 57<br>(11.80) | 27<br>(15.34) | 16<br>(13.56) | 5 (15.15) | 25<br>(10.16) | **<0.001** |
| Swollen tongue | 25 (1.60) | 20 (4.26) | 7 (4.14) | 6 (5.17) | 1 (3.13) | 11 (4.56) | **0.001** |
| **Neurological manifestations, n (%)** |  |  |  |  |  |  |  |
| Non-migraine headache | 225 (21.55) | 98<br>(31.82) | 21<br>(22.34) | 14<br>(21.21) | 4 (21.05) | 71<br>(39.89) | **<0.001** |
| Migraine headache | 218 (20.62) | 79<br>(25.82) | 25<br>(23.15) | 24<br>(29.27) | 8 (36.36) | 37<br>(23.57) | 0.053 |
| **Hematological manifestations, n (%)** |  |  |  |  |  |  |  |
| IDA | 1208 (55.26) | 381<br>(62.98) | 140<br>(65.73) | 93<br>(63.27) | 26<br>(66.67) | 182<br>(60.67) | **0.001** |
| Thalassemia | 120 (7.57) | 51<br>(10.87) | 14 (8.54) | 9 (7.96) | 4 (11.43) | 28<br>(11.67) | **0.023** |

**Abbreviations:** DH, dermatitis herpetiformis; IDA, iron deficiency anemia; N, number.

### Extra-intestinal manifestations

Celiac patients with cutaneous manifestations had a significantly higher prevalence of mouth aphthae (30.9% *vs.* 21.4%, respectively; *p*<0.001), mouth ulcers (11.8% vs. 6.3%, respectively; *p*<0.001), and swollen tongue (4.2% vs. 1.6%, respectively; *p*=0.001). The prevalence of non-migraine headaches was significantly higher among patients with cutaneous disorders than celiac patients without skin manifestations (31.8% *vs.* 21.5%, respectively; *p*<0.001). The prevalence of both types of anemia was significantly higher among celiac patients with skin manifestations (62.9% *vs.* 55.2%; *p*=0.001 for IDA and 10.8% *vs.* 7.5%; *p*=0.023 for thalassemia, respectively) (**Table 2**).

### Associated disorders

Celiac patients without cutaneous manifestations showed a higher prevalence of type I DM (14.4%) compared to patients with skin disorders (6.9%) which revealed a statistically significant difference (*p*<0.001). While there was a significantly higher prevalence of osteoarthritis (OA) (3.4% *vs.* 1.1%, respectively; *p*=0.001) and Sjögren Syndrome (0.8% *vs.* 0.2%, respectively; *p*<0.001) among patients with cutaneous disorders compared to celiac patients without skin manifestations, the prevalence of other associated disorders was similar between the two groups (*p*>0.05) (**Table 3**).

**Table 3.** Prevalence of associated disorders and family history of associated diseases in patients with and without cutaneous manifestations.

| Variables | Patients without cutaneous manifestations (n= 3564) | Patients with cutaneous manifestations (n=793) |  |  |  |  | P-value |
| --- | --- | --- | --- | --- | --- | --- | --- |
|  |  | Total (n=793) | Eczema (n=251) | DH (n=179) | Psoriasis (n=47) | Others (n=423) |  |
| Associated disorders, n (%) |  |  |  |  |  |  |  |
| Hypothyroidism | 368<br>(19.91) | 125<br>(22.85) | 37<br>(20.00) | 23 (17.04) | 6 (16.67) | 74<br>(25.96) | 0.135 |
| Hyperthyroidism | 49 (2.90) | 19 (3.81) | 5 (2.86) | 2 (1.57) | 1 (3.03) | 14 (5.47) | 0.305 |
| Type I DM | 265<br>(14.48) | 36 (6.99) | 15 (8.43) | 5 (3.76) | 1 (2.70) | 19 (7.28) | **<0.001** |
| Type II DM | 29 (1.72) | 15 (3.01) | 4 (2.30) | 3 (2.38) | 2 (5.56) | 8 (3.15) | 0.073 |
| RA | 52 (3.34) | 31 (6.57) | 12 (7.79) | 6 (4.92) | 3 (10.34) | 16 (6.53) | 0.599 |
| Growth impairment | 51 (3.60) | 14 (3.45) | 4 (2.78) | 3 (2.91) | 0 (0.00) | 8 (3.83) | 0.143 |
| OA | 17 (1.11) | 15 (3.41) | 7 (4.46) | 3 (2.83) | 2 (6.45) | 6 (2.61) | **0.001** |
| Down syndrome | 20 (2.42) | 10 (4.48) | 3 (4.00) | 2 (3.33) | 2 (11.11) | 4 (3.51) | 0.565 |
| AI hepatitis | 12 (0.87) | 2 (0.50) | 2 (1.37) | 0 (0.00) | 0 (0.00) | 0 (0.00) | 0.456 |
| Turner syndrome | 10 (0.67) | 2 (0.45) | 0 (0.00) | 1 (0.88) | 0 (0.00) | 1 (0.43) | 0.732 |
| Sjögren syndrome | 3 (0.20) | 4 (0.88) | 2 (1.33) | 1 (0.87) | 0 (0.00) | 2 (0.84) | **<0.001** |
| Infertility | 2 (0.13) | 1 (0.22) | 0 (0.00) | 0 (0.00) | 0 (0.00) | 1 (0.42) | 0.883 |
| **Family history of associated disorders, n (%)** |  |  |  |  |  |  |  |
| CD | 268<br>(16.37) | 96 (20.17) | 52<br>(33.33) | 35 (28.93) | 5 (17.24) | 33<br>(13.25) | 0.054 |
| Hypothyroidism | 359<br>(21.29) | 124<br>(24.90) | 40<br>(24.54) | 24 (19.83) | 4 (13.33) | 80<br>(30.42) | 0.088 |
| Hyperthyroidism | 83 (5.32) | 21 (4.56) | 5 (3.36) | 4 (3.42) | 1 (3.57) | 12 (4.94) | 0.514 |
| Type I DM | 131 (8.29) | 50 (10.68) | 12 (8.00) | 7 (5.88) | 3 (10.34) | 36<br>(14.63) | 0.109 |
| Type II DM | 197<br>(12.24) | 66 (13.75) | 15 (9.68) | 13 (10.92) | 6 (20.00) | 40<br>(15.75) | 0.380 |
| RA | 52 (3.34) | 31 (6.57) | 12 (7.79) | 6 (4.92) | 3 (10.34) | 16 (6.53) | **0.002** |
| Skin disorders | 16 (1.86) | 24 (10.39) | 17<br>(20.00) | 12 (18.75) | 0 (0.00) | 8 (6.78) | **<0.001** |
| Liver disorders | 20 (2.42) | 10 (4.48) | 3 (4.00) | 2 (3.33) | 2 (11.11) | 4 (3.51) | 0.102 |
| Down syndrome | 10 (0.67) | 2 (0.45) | 0 (0.00) | 1 (0.88) | 0 (0.00) | 1 (0.43) | 0.599 |
| Hashimoto's thyroiditis | 6 (0.40) | 2 (0.44) | 0 (0.00) | 0 (0.00) | 0 (0.00) | 2 (0.84) | 0.896 |
| Sjögren syndrome | 3 (0.20) | 4 (0.88) | 2 (1.33) | 1 (0.87) | 0 (0.00) | 2 (0.84) | **0.033** |
| Thalassemia | 6 (0.40) | 0 (0.00) | 0 (0.00) | 0 (0.00) | 0 (0.00) | 0 (0.00) | 0.178 |
| Turner syndrome | 2 (0.13) | 1 (0.22) | 0 (0.00) | 0 (0.00) | 0 (0.00) | 1 (0.42) | 0.673 |

**Abbreviations:** AI, autoimmune; CD, celiac disease; DH, dermatitis herpetiformis; DM, diabetes mellitus; N, number; OA, osteoarthritis; RA, rheumatoid arthritis.

### Family history of associated disorders

The prevalence of a positive FH for rheumatoid arthritis (RA) (6.5% *vs.* 3.3%; *p*=0.002), skin disorders (10.3% *vs.* 1.8%; *p*<0.001), and Sjögren syndrome (0.8% *vs.* 0.2%, respectively; *p*=0.033) was significantly higher among celiac patients with cutaneous manifestations than those without skin disorders. There was no statistically significant difference regarding a positive FH for liver diseases, Down syndrome, Hashimoto’s thyroiditis, Sjögren syndrome, thalassemia, and Turner syndrome between the study groups (*p*>0.05) (**Table 3**).

### Multivariate logistic analysis

The multivariate logistic analysis of demographic and clinical characteristics revealed that among gastrointestinal manifestations, having constipation was an independent factor for developing cutaneous manifestation (OR=1.62, 95% CI 1.10-2.38, *p*=0.01). Regarding oro-dental manifestations, CD patients with mouth aphthae had a 3.10 times greater likelihood (95% CI 1.80-5.32, *p*=0.00) of developing skin disorders. IDA (OR=1.54, 95% CI 1.05-2.24, *p*=0.02) and thalassemia (OR=2.78, 95% CI 1.19-6.50, *p*=0.01) significantly increased the likelihood of skin disease development. On the other hand, type I DM was identified as a protective factor for skin manifestation development (OR=0.20, 95% CI 0.05-0.81, *p*=0.02), while OA (OR=6.15, 95% CI 1.15-32.78, *p*=0.03) and Sjögren syndrome (OR=4.82, 95% CI 1.98-11.73, *p*=0.00) were found to be independent risk factors for developing cutaneous manifestations. Regarding the FH of associated disorders, it was found that a positive FH of skin disorders was associated with a 7.99 times greater likelihood (95% CI 2.44-26.13, *p*=0.00) of developing cutaneous disorders. The results of multivariate logistic analysis are presented in **Table 4**.

**Table 4.**
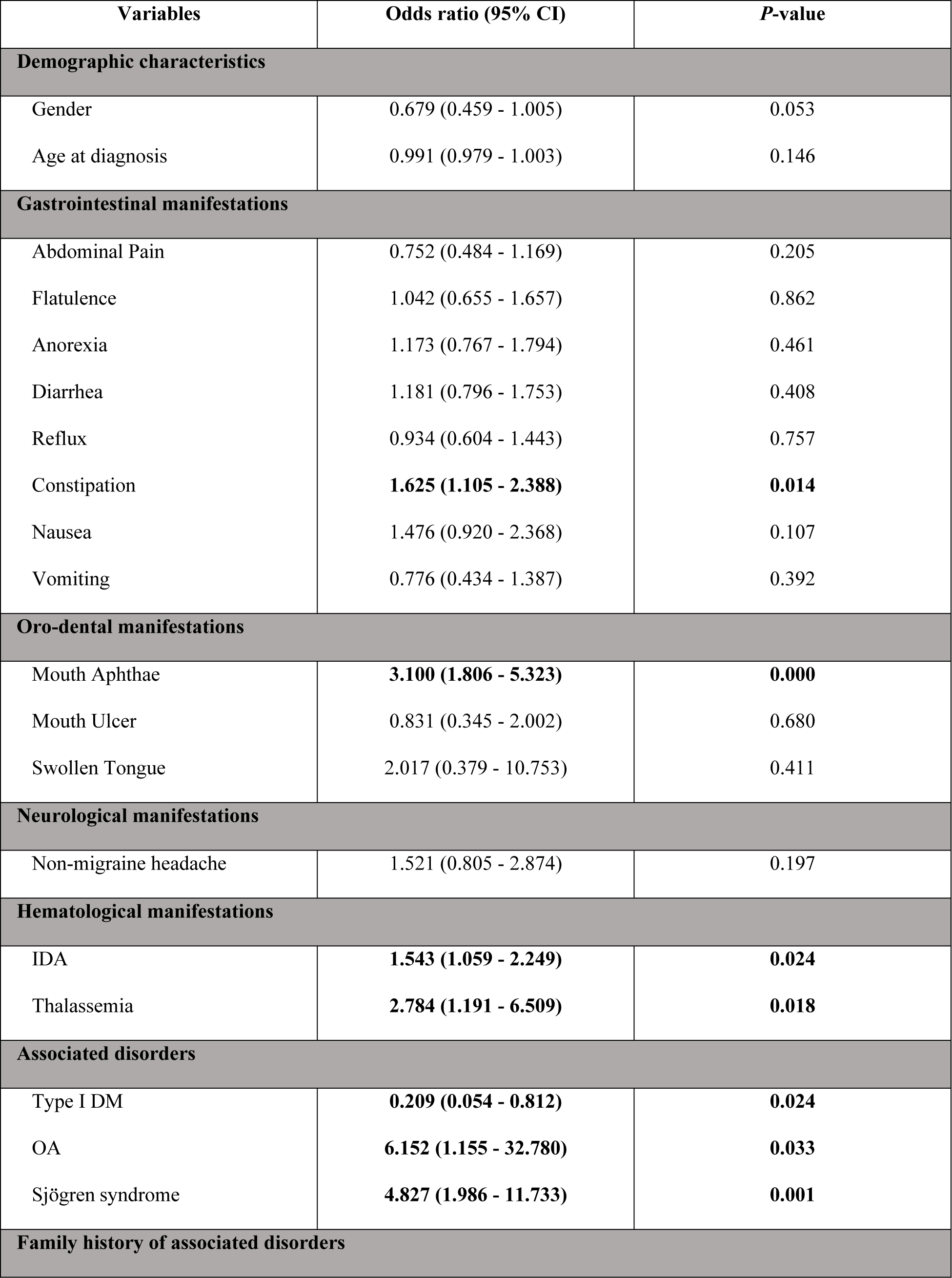
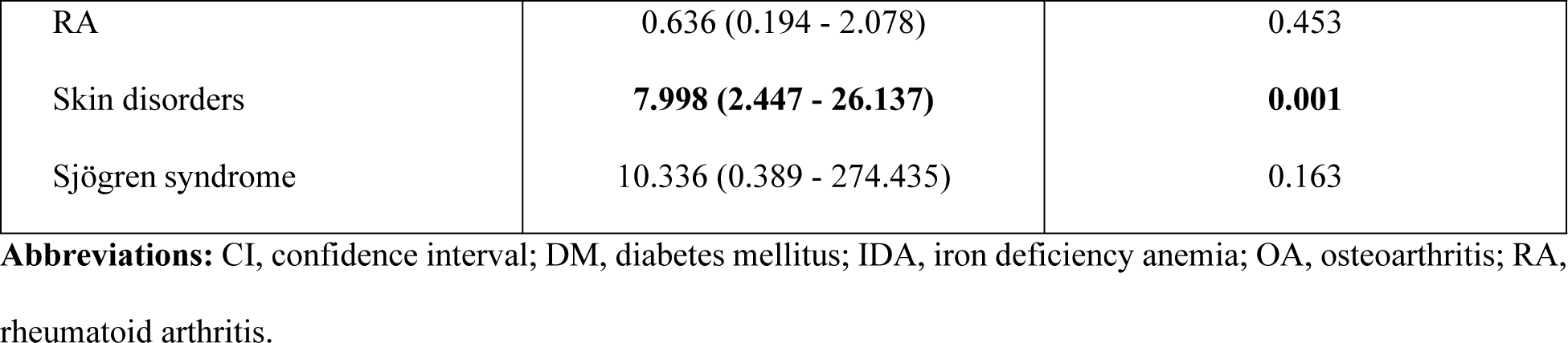
Multivariate logistic analysis of the association between demographic and clinical characteristics and development of skin disorders.

## Discussion

Although for many years it was believed that CD mostly presents with typical gastrointestinal manifestations, in recent years, a growing body of literature has shown that CD clinical presentation has dramatically changed. A considerable proportion of CD cases now present with extraintestinal manifestations, with certain cases demonstrating such manifestations prior to the onset of gastrointestinal manifestations, and in some instances, serving as the sole clinical presentation of the underlying disease. Cutaneous disorders are considered among the most common extraintestinal manifestations in patient with CD. Thus, understanding the cutaneous manifestations of CD is crucial for early diagnosis, timely initiation of treatment, and better disease management, as well as managing concurrent disorders and related manifestations. The findings revealed that 18.2% of CD patients had cutaneous manifestations, with eczema being the most prevalent skin disorder, followed by DH and psoriasis with a prevalence of 3.7%, 3.4%, and 2.9%, respectively. The study identified constipation, mouth aphthae, underlying IDA, thalassemia, OA, Sjögren syndrome, and a positive FH of skin disorders as independent risk factors, while underlying type 1 DM was a protective factor for the development of cutaneous manifestation in CD patients. However, it is noteworthy that although this study found these factors to be independently associated with cutaneous disorders, no causal relationship can be established between the aforementioned factors and the development of skin diseases.

In our study, a total of 18.2% of patients revealed cutaneous disorders, with eczema being the most common skin disease, followed by DH, and psoriasis. Several studies have shown an association between CD and various skin disorders. Niknam et al. reported the prevalence of cutaneous manifestations ranging from 10.9% in children to 27.7% in adult patients with CD (20). Salarian et al. conducted a cross-sectional study demonstrating that eczema was the most common cutaneous manifestation among pediatric and adolescent patients with CD, with a prevalence of 18.6%, nearly 5 times higher than eczema prevalence in our study (21). Additionally, we identified DH as the second most common skin disorder among CD patients, with a prevalence of 3.4%, while Lebwohl et al. reported DH in only 0.9% of celiac patients (8). These differences between studies may be attributed to variations in the prevalence of cutaneous disorders across different countries, influenced by geographical region, race, and genetic background. Differences in study design, sample size, and diagnostic methods could also account for discrepancies among the studies.

Our study revealed that CD patients with constipation had a 1.6-fold greater likelihood of developing cutaneous manifestations. Although the exact underlying mechanisms linking gastrointestinal disorders and skin diseases are not fully understood, intestinal microbiome, metabolic, and neuroendocrine interactions between the gut and skin have been proposed to be accountable for this association (22-27). Our study also found that mouth aphthae was associated with a 3.1-fold higher likelihood of developing cutaneous disorders in CD patients. Previous studies have indicated that patients with various dermatological diseases may also exhibit oro-dental manifestations. For instance, a recent study in Saudi Arabia showed that 14.5% of patients with cutaneous disorders had various oral manifestations, including perioral dermatitis, melanotic macule, mucocele, etc. (28). Altogether, given the higher likelihood of skin disorders in CD patients with constipation or mouth aphthae manifestations, clinicians should frequently monitor these patients for any skin manifestation.

Regarding the underlying disorders, the present study revealed that the likelihood of developing skin disorders was 1.5- and 2.7-fold higher in patients with IDA and thalassemia, respectively. Studies have shown that several main components of skin tissue, which play important roles in maintaining normal skin structure and function, contain iron (29). Therefore, IDA may predispose affected patients to the development of various skin disorders. Furthermore, studies have indicated that patients with thalassemia may also present with concurrent cutaneous manifestations due to several reasons including iron overload, sun exposure, copper deficiency, skin dehydration, inappropriate histamine release, and hemosiderosis (30-32). Our study also highlighted that OA and Sjögren syndrome were associated with a 6.1- and 4.8-fold greater likelihood of developing cutaneous manifestations in CD patients, while the likelihood of skin disease development in CD patients with Type I DM was 0.2-times lower. Studies have demonstrated that several AI disorders can co-exist and having one AI disease may increase the risk of developing others (33-35). Therefore, in patients with AI disorders, there may be a higher chance of developing immune-mediated skin disorders, as observed in our study. Although it has been demonstrated that DM is associated with various types of skin disorders (36), some studies have revealed that receiving antidiabetic treatments and appropriate glycemic control may reduce the risk of developing cutaneous manifestations (37-39). Thus, the inverse relationship between Type I DM and the risk of developing skin disorders found in our study might be mediated by clinical conditions of diabetic patients regarding reception of antidiabetic agents and glycemic control status. Nevertheless, the inverse association between Type I DM and the likelihood of developing skin disorders observed in our study warrants further evaluation.

In addition, regarding the FH of associated diseases, CD patients with a positive history of skin disorders were at a 7.99-fold higher risk for the development of cutaneous manifestations. In accordance with our results, previous studies also reported that a positive FH of skin disorders can influence the onset, course, and presentation of skin diseases (40-42). Altogether, it is crucial for clinicians to be aware of the most common cutaneous manifestations in patients with CD, especially those with constipation, mouth aphthae, IDA, thalassemia, OA, Sjögren syndrome, and a positive FH of skin diseases, and regularly assess these patients for the development of any skin presentations, as early diagnosis and treatment of underlying skin disorders may be beneficial in improving CD prognosis and clinical outcomes. Moreover, although no routine screening protocol for early detection and diagnosis of skin diseases in patients with a FH of cutaneous disorders has been proposed, these individuals can undergo regular clinical examination with the purpose of detecting any cutaneous presentations.

The current study has several strengths. Firstly, it boasts a relatively large sample size consisting of more than 4300 confirmed cases of CD. Secondly, a large variety of clinical variables were evaluated. Nonetheless, the study also has several limitations which need to be acknowledged. First, since the study was retrospective in nature and relied on self-reported data, some degrees of recall bias might be introduced. Second, the present study was conducted in a single center which resulted in a less diverse sample and decrease the generalizability of findings. Third, detailed information regarding the specific diagnostic criteria used for identifying cutaneous manifestations was not provided in the study, therefore, it would be difficult to assess the reliability and validity of the current findings. Fourth, data regarding the duration of CD was not assessed in our study, which might affect the risk of developing skin manifestations. Thus, further cohort studies with large sample size and long follow-up duration are needed to find the risk and protective factors of skin disorder development in patients with CD. In addition, experimental studies are warranted to comprehend the pathophysiological mechanisms underpinning the association between CD and cutaneous disorders.

## Conclusion

The present study revealed that approximately 18.2% of patients with CD may experience various cutaneous manifestations and the likelihood of developing skin disorders seems to be higher in CD patients with constipation, mouth aphthae, IDA, thalassemia, OA, Sjögren Syndrome, and a positive FH of cutaneous disorders. Given the significant prevalence of skin disorders in CD patients, especially those with aforementioned clinical characteristics, it is critical for clinicians to be aware of common cutaneous manifestations among these patients to make a timely diagnosis and early initiation of treatment for both the underlying CD and any concurrent cutaneous disorder, ultimately leading to improved disease prognosis, decreased disease severity and burden, and increased patient quality of life. Further studies are needed to evaluate possible causal relationships between the aforementioned conditions and development of cutaneous disorders. Moreover, it could be also beneficial to evaluate the impact of implementing a GFD on the clinical condition of CD patients with cutaneous manifestations. Additionally, the potential effect of CD duration and severity on risk of developing various skin manifestations should be further studied.

## Data Availability

All relevant data are within the manuscript and its Supporting Information files.

## Acknowledgment

None.

